# Insurance as an instrument of financial security in addressing mental illness among agricultural workers in the LMICs

**DOI:** 10.1101/2022.06.09.22275910

**Authors:** Sandip K. Agarwal, Snehil Gupta, Vijender Singh, Roshan Sutar, Drupad Nair

## Abstract

**Introduction:** Agriculture is a high-risk occupation globally, with risk intensities being higher in low-and middle-income countries (LMICs). Formal risk-mitigation instruments are absent in LMICs. Prevalence of financial insecurity often acts as a prominent stressor predisposing to various mental illnesses among the agrarian workers in LMICs.

**Aims:** We review the existing scientific evidence from LMICs on the role of insurance in improving the mental health of rural workers. Our research identifies the different insurance interventions available for agrarian or rural workers in LMICs, and review their effectiveness - overall and across sub-groups in preventing the development of mental illnesses or reducing existing ones.

**Methodology:** Our review included both peer and non-peer-reviewed literature. We involved people with lived experience (PWLE) that included farmers, workers, NGOs and health workers, policy researchers etc. with experiences from India, Bangladesh, Nepal, Peru, and South Africa. Inputs from PWLE helped in defining our key concepts for the study and in curating a list of keywords for literature search. We identified 79 articles of which we reviewed 47 articles that reported effectiveness of financial interventions, primarily Cash transfer (CT), Pension and Health insurance, Workfare and Microfinance on the mental health of rural workers in LMICs. A cash transfer (CT) is a direct transfer payment of money to an eligible person. Unlike Unconditional CT (UCT), conditional CT (CCT) are conditional upon completing specific actions beyond being eligible for CT such as sending children to school or making regular health visits. Microfinance is provision of banking service to individuals or groups who otherwise would have no access to financial services. Workfare program require participants to perform usually public-service work to receive payment.

**Insights from PWLE:** We learned from PWLE that it is hard to distinguish between self and wage employment as workers are engaged in multiple livelihood activities, and similarly between rural and urban workers due to seasonal migration. Workers continue to work in old age as there is no concept of retirement. PWLE reported that public assistance program whether they supplement income or consumption are beneficial. Access to low cost credit enhances financial security as most workers operate in highly credit-constrained environments.

**Key findings:** (i) Pension and health insurance led to a significant reduction in symptoms of depression and anxiety among workers, particularly among the elderly. (ii) Workfare participation led to a reduction in depression among women by increasing income security. However, in addition to financial security, non-pecuniary benefits of employment were also observed among the unemployed refugee men. (iii) CT led to a reduction in suicides among farmers during adverse income shocks, and in general improved mental health of recipients. However, when the recipients perceived CT as stigmatizing or perceived the compliance condition (as in CCT) as an additional burden, the effects of CT on mental health were negative. (iv) Microfinance schemes had mixed effects on mental health of the participants, primarily women. While it led to a reduction in depression and anxiety, loan repayment was often reported to be stressful.

**Recommendation for practice:** Mental health should be incorporated as an additional welfare parameter in the cost-benefit analysis of policy evaluation as evidence suggest that higher income or consumption do not necessarily improve mental health. While pension and health insurance can bring in positive changes in mental health of workers, CT and microfinance schemes are no silver bullets for improving psychological well-being. CT and microfinance interventions can have an adverse effect on mental health of recipients which depend upon their design and delivery.

**Recommendation for research:** We believe that lack of mental health data largely inhibits evidence-based research on mental health. For example, we did not find any study that evaluated agricultural insurance or price support scheme in spite of these having existed in LMICs for some time now. Evaluation of existing insurance interventions on parameters of mental health is only possible if data collection on mental health variables are encouraged.

## Introduction and Background

Agricultural livelihoods are impacted by several risks which include production-, market-, institutional-, idiosyncratic-, and financial-risks [1–3]. While fundamentally similar, the intensity of these risk is higher in Low and Middle-Income countries (LMICs) due to weak institutions, and an increased vulnerability to disease hazards and pest infestation in tropical climates [1]. More than 70% of the farms in LMICs are less than 2 hectares in size, with a trend of increasing numbers of farms and smaller farm size, particularly in South Asia [4, 5]. LMICs have a predominantly rural and agrarian population with a highly informal agricultural sector, with an overwhelming proportion of agrarian households living in poverty.

Recent scientific evidence has found bidirectional causality between poverty and mental illness, suggesting a poverty trap [6]. With LMICs housing more than 80% of the global population affected by mental health disorders, vulnerability of rural poor to these disorders is particularly high [7]. Multiple sources of evidence of mental illness and suicide exist among agriculture workers, linked to the usage (acute or chronic) of chemicals and pesticides on the farm [8–12]. Climate change has exacerbated the incidence of mental illness and suicide among agricultural workers [13–17]. Additionally, strong evidence exists identifying socio-economic and financial factors as key predictors of mental illness and suicide among the agrarian population in LMICs [18, 19].

Agricultural sectors in LMICs are highly informal, therefore formal or market-based instruments to mitigate risk are generally non-existent. Furthermore, low agricultural incomes, and high income volatility aggravates the already poor mental health of agrarian workers. Among many, financial burden and farm losses have been one of the crucial predictors of poor mental health and cases of suicide among the agrarian workforce [18, 20–30]. Therefore, we hypothesize that insurance can have a significant role in providing adequate protection to agricultural workers in adverse income situations, which can subsequently prevent development of mental illness or further deterioration of mental health conditions of the agrarian workers.

Our definition of insurance is broad, and includes financial interventions that potentially enhance financial security of rural workers by reducing income uncertainty. Our concept of insurance includes both market-based insurance instruments and social protection-based welfare programs for which the beneficiaries need not contribute to be eligible for the benefits or their contribution for participating in the program is often highly subsidized.

We attempt to answer the following questions by reviewing the existing evidence:

1. What are the different insurance interventions available for agrarian or rural workers in LMICs?
2. How effective are existing insurance interventions in preventing the development of mental illnesses or reducing existing ones?
3. What is the evidence of heterogeneity in effectiveness of available insurance interventions across sub-populations?

## Methodology

Our study methodology included a mix of review of peer reviewed literature from academic journals and non-peer reviewed gray literature. We actively involved people with lived experience (PWLE) by using inputs from their interviews and focus group discussions to inform our research design.

Input from PWLE was used for two purposes. Firstly, it helped us to improve our research design by aligning our definitions with the experiences of PWLE. Secondly, input from PWLE was used to curate a list of keywords used for literature search. The diversity of the PWLE engaged in our study is provided in table 1. A total of 39 distinct PWLE were involved, of which around 30% were women and 66% were from India, including regions with high agrarian distress like Mandya, Karnataka; Vidarbha, Maharashtra; Punjab; and Sundarbans in West Bengal. While the Sundarbans bear an unproportionate burden of climate change in the form of intermittent cyclones and floods, other regions have high incidence of agrarian suicide and drug and alcohol abuse.

**Table 1.** PWLE across diverse occupational experience and gender (male (M) and female (F))

| Countries | India |  | Bangladesh |  | Nepal |  | Peru & South Africa |  |
| --- | --- | --- | --- | --- | --- | --- | --- | --- |
| PWLE Occupation / Gender | M | F | M | F | M | F | M | F |
| Farmers and Farm workers | 11 | 1 | 2 | 2 | 2 | 1 | 0 | 0 |
| Rural Credit institution | 2 | 0 | 0 | 0 | 0 | 0 | 0 | 0 |
| Farmer Producer Organization | 2 | 0 | 0 | 0 | 0 | 0 | 0 | 0 |
| Social and Health Workers | 1 | 1 | 0 | 0 | 0 | 0 | 0 | 0 |
| NGOs | 1 | 3 | 0 | 0 | 0 | 0 | 0 | 0 |
| Researchers (including Policy Think Tanks) | 2 | 2 | 2 | 2 | 1 | 0 | 1 | 0 |

Our key definitions evolved during the study with the involvement of PWLE, as we continuously updated our definitions to reflect the understanding of PWLE rather than what we had initially conceived. Our definition of agrarian workers relied upon a recent collaborative report by the International Labor Organization (ILO) and Food and Agriculture Organization (FAO) on extending social protection to rural populations [31]. Agricultural workers were defined as “predominantly rural, engaged in wage or self-employment who often work in temporary employment (seasonal or casual) and part-time employment.” It is often true that in informal work settings the definition of a workplace is blurred. We learned from our PWLE that the distinction between self and wage employment is blurred too, and so is the distinction between rural and urban workers, as rural workers often migrate to urban areas for work during lean season, while they continue to remain engaged in agriculture during the farming season. Similarly, workers in rural areas are simultaneously engaged in multiple livelihood options, which is an informal way of risk diversification. Based on these discoveries, we modified our unit of analysis from workplace to workers which included migrant workers.

Our definition of insurance included interventions that enhance financial security of rural workers. We included social insurance and assistance schemes for the rural workers in our review following the definition of social protection in the ILO and FAO joint report. We also learned from PWLE that eligible beneficiaries for social protection are identified based on socio- economic conditions rather than sector of employment. Hence, income threshold is used as a key parameter in targeting the beneficiaries. However, PWLE mentioned that if there is some form of insurance, the method of targeting is inconsequential. Considering the above, we expanded our research to include welfare programs too, and included rural households since households included working members too. We also included social pension insurance as an intervention, as PWLE informed us that workers continue to work in their old age, because there is no concept of retirement in the informal sector. We also learned that in credit constrained environments, access to formal credit provides financial security and reduces their worries. Subsequently, we included credit interventions too.

Mental illnesses for our review primarily included depression, anxiety, stress, and suicide. We also included other mental health issues, such as entrenched negative beliefs, ruminations, and suicidal ideation. While we did not include studies that exclusively focused on quality of life, subjective well-being, or level of disability, we included studies that link either of these with one of our primary variables above.

A second purpose of the inputs from PWLE was to identify the keywords that were used for literature search. Array A2 in the appendix provides the master list of the keywords that were created from interactions with PWLE. Prior to finding the literature, relevant databases (as in table A1 in the appendix) were shortlisted from the list of database and search engines on Wikipedia.

The research design and the search strategy used to identify the relevant literature have been depicted in Figure 1.

**Figure 1.**
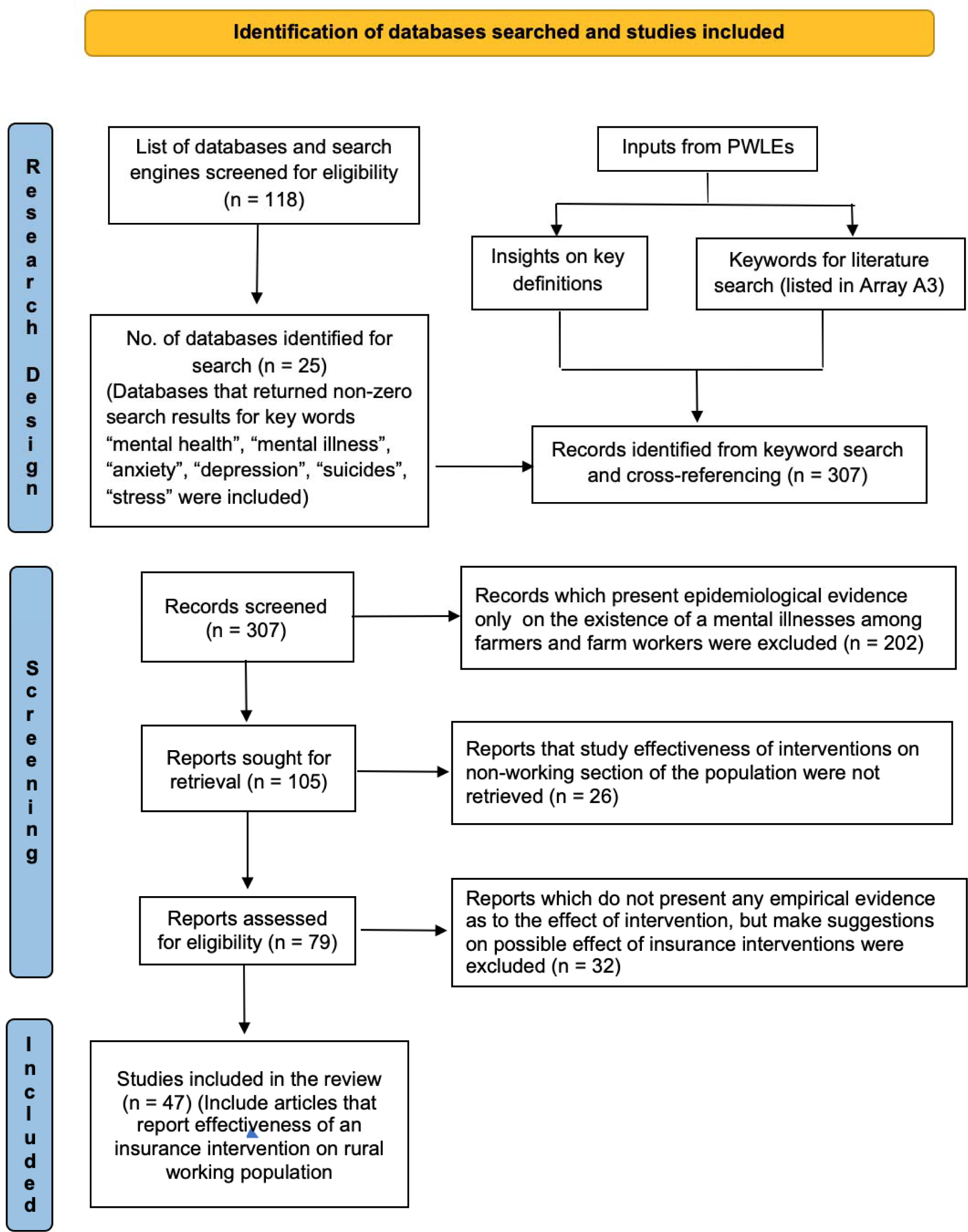
Flowchart of research design and articles included for review

## Results

Our final review included 47 articles that reported effectiveness of financial interventions on the rural or agrarian working population in LMICs. Table A3 in the appendix lists a detailed summary table of these 47. Additionally, we also have another 32 articles that make suggestions and claims about several financial interventions that can address agrarian distress, where farmer suicides are a manifestation of agrarian distress. These studies are rather suggestions and opinions unlike the 47 studies we have included in our review, which provide robust empirical evidence of the effectiveness of insurance interventions. We provide a brief review of ideas and summary of these articles in section A4 and table A5 in the appendix.

The primary financial interventions that the 47 articles span for our review include Cash transfers, Pension insurance, Health insurance, Workfare, Microfinance and some miscellaneous but innovative interventions.

### **(a)** Cash Transfers (CT)

A cash transfer (CT) is a direct transfer payment of money to an eligible person. Unconditional cash transfers (UCTs) are given to beneficiaries without any specific requirements beyond eligibility, whereas conditional cash transfers (CCTs) are conditional upon completing specific actions such as sending children to school or making regular health visits.

Studies have found causal effects of both UCTs and CCTs on psychological well-being of the beneficiaries by a reduction in at least one of the following mental health variables - depression, stress, anxiety, and suicide in rural settings in LMICs [32–39]. Haushofer and Shapiro (2018) found that the positive effect of CT on psychological outcomes persisted in the long term as well. In a comparative study of CT and psychotherapy, Haushofer et al. (2020a), found CT to be more effective than psychotherapy in reducing stress and depression. However, another study by the same authors did not find CT to be effective in reducing stress among informal workers in Kenya working in hazardous conditions with minimal safety equipment [40].

The mental health effects of CT also varied across sub-population. For instance, Christian et al. (2019) found CT to be more effective in reducing suicide among farmers by mitigating the depressive symptoms resulting from negative income shock due to poor rainfall that lowered agricultural productivity. Kilburn et al. (2016) reported a reduction in depression following a CT scheme for Orphans and Vulnerable Children in Kenya. They found a positive effect of CT on males aged 15-24yrs, but the effect was non-significant for female counterparts. Yet another study based on CCT (Prencipe et al., 2021), involving youth aged 14-28yrs, found a reduction in depressive symptoms among the males, however, symptoms worsened among females.

Notably, CT programs can also adversely affect a recipient’s mental health, if stigma and shame are associated with the receipt of the CT, as in the case of China’s Dibao, which is an income assistance program for the ultra-poor [41, 42]. However, the above results as in Wu et al. (2021), establishes only correlation, not causality. As Dibao beneficiaries are already an ultra-poor population, it is likely that their baseline mental health was worse (compared to the non-recipients) and the intervention likely had a positive effect.

Another apparently similar interventions are cash grants, which are CTs offered with the expectation of providing opportunities to rural youth for self-employment in the non-farm sector. No significant effect on the level of depression or distress among the grant recipients whether in the short-term or long-term basis were observed [43–45]. All the above studies measured depression and distress using a pre-validated instrument that was developed locally in the conflict and violence ridden settings of rural Northern Uganda [46].

### **(b)** Pension Insurance

Old age pension is an allowance paid to elderly people by the government or their employers after a certain age when they retire and stop working. While there is no concept of old age retirement in the informal sector as people continue to work in their old age, pension insurance provides elderly people with income security. Under non-contributory pensions, the entire pension insurance scheme is financed by the government, whereas participants have to partly contribute to the pension fund in a contributory pension.

Non-contributory pensions have been found to have a profound impact on the beneficiaries through a significant reduction in their depression, but concern exists about their long-term positive effects [47–51]. Similarly, contributory social pensions have been shown to be associated with lower depression among the elderly rural recipients [52–55]. While another study, involving marginalized population in South Africa, reports a non-significant association between pension and anxiety and depression symptoms [57].

The heterogeneous effects of contributory social insurance have also been observed across sub-population. The largest reduction in depression was observed among the least educated and the lowest income elderly populace [52, 55]. Additionally, Wang and Zheng (2021) found no effects of pension on depression among the elderly males relative to females; a reduction in depression among the elders were found who lived independently vs. those who lived in extended households.

### **(c)** Health Insurance

Health insurance covers the cost of medical care for an individual. Often, people without health insurance suffer from poor health as they are unable to access medical care and treatment for themselves and their families. As health insurance is absent in informal sector employment, governments in many LMICs have introduced floated social health insurance to provide health coverage for their population.

In the studies we reviewed, social health insurance participation significantly reduced depression [58–61]. The above studies evaluated the causal effect of subsidized contributory social health insurance in China. Catastrophic medical insurance was also found to be associated with lower reduced depression and anxiety among the elderly [62, 63]. Non-significant effects also exist for the effects of social health insurance on psychological well-being [53]. In an experimental study in Kenya, free health insurance for urban workers working in hazardous conditions reduced stress compared to an equivalent amount of CT, which was inconsequential to mental health [40].

Differential effects of health insurance across sub-populations has also been reported. For instance, Wang et al. (2009) found the largest effect of the intervention on people older than 55 years and those who were already ill prior to the intervention. Similarly, for Sun and Lyu (2020) the largest positive effect of the intervention was on lower income individuals and for Sun et al. (2021) it was most effective for males and population below 65 years in a sample population of 45 years and above.

### **(d)** Workfare

Workfare is a welfare program in which recipients are required to perform usually public-service work to receive payment. Workfare can reduce symptoms of depression and anxiety [38,64,65]. Ravi and Engler (2015) and Tsaneva & Balakrishnan (2019) used the National Rural Employment Guarantee Scheme (NREGS) in India as the workfare intervention for their study while Hussam et al. (2021) used a workfare and CT experiment in the Rohingyas refugee camp in Bangladesh. Hussam et al. (2021) found higher reduction in symptoms of depression and stress for the workfare intervention relative to UCT among the males; no significant difference was observed among the females. Bhanot et al. (2018) found similar results of significant positive impacts of workfare on psychological well-being, relative to those who got the same benefit by waiting instead of working. Pailler and Tsaneva (2018) found adverse effects of climate change on depression, sleep and cognitive difficulties in rural agrarian populations in India, which they suggest can be mitigated by a workfare program like NREGS.

### **(e)** Microfinance

Microfinance or microcredit is a type of banking service provided to unemployed or low-income individuals or groups who otherwise would have no other access to financial services. Microcredit participation can improve mental health [68–69]. For instance, Mohindra et al. (2008) found that microcredit participation reduces emotional stress among agrarian women participants in a South Indian village. Similarly, Hamad and Fernald (2015) found that microcredit participation reduces depression among Peruvian women. Both the studies also report higher effectiveness of microcredit participation among women who had longer duration of microcredit participation.

Evidence of the ineffectiveness of microcredit participation or its adverse effects on mental health also exist [70, 72–74]. Participation in a group-based savings and loan scheme did not have any significant reduction in depression, anxiety, and post-traumatic stress disorder (PTSD) among women survivors of sexual violence in Eastern Democratic Republic of Congo (DRC). However, in the same context, another variant of microfinance where livestock instead of money were loaned was effective in reducing depression and PTSD among women [71]. Research from Bangladesh has also found microcredit participation to be associated with higher emotional stress [72, 73]. In a study of marginally rejected microloan borrowers in South Africa, who were extended loan on second consideration, it was found that access to microloan increased stress but reduced depression [74]. Similarly, in a study among HIV positive Kenyan farmers, Hatcher et al. (2020) have found that participants of a multi-sectoral intervention with a limited liability loan reported higher levels of stress associated with loan repayment even though the qualitative data report reduction in symptoms of depression and anxiety.

### **(f)** Miscellaneous

Liang et al. (2014) evaluated the effectiveness of social security policies towards rehabilitation of landless farmers and found them to be associated with lower levels of depression and anxiety. The policies included land acquisition compensation, medical insurance, pension, employment security, basic livelihood guarantee and housing compensation.

Living wage (i.e., a wage significantly higher than the minimum wage) in combination with significant workplace improvements was found to be associated with lower scores of depressions among apparel factory workers in the Dominican Republic [77].

An intervention of affordable low-cost daycare programs, targeted towards poor women in rural India, was associated with a modest reduction in mental distress [78].

In a multi-country, “Graduation” program which consisted of a holistic set of services that included the granting of a productive asset, a consumption support, a savings account, and training support, did not have a consistent effect on mental health in all six countries [79].

## Discussion

We provide a synthesis of results from the previous section to build insights on the effectiveness of insurance on the rural (mostly agrarian) population in LMICs.

The CT has been the most widely investigated intervention. The evidence on the effectiveness of CT had been very diverse in different context and across different sub-population. While most evidence point towards positive effects of CT in improving mental health of the beneficiaries, negative effect also exist. Similarly, microcredit is another intervention for which we find conflicting evidence on its effect on mental health of recipients. For interventions that include health and pension insurance and workfare, overall evidence indicates toward their positive effect on the mental health of beneficiaries.

A crude comparison and synthesis of results from our review help us make some compelling inferences. Firstly, it is likely that the effectiveness of a particular intervention depends largely upon the cause and the nature of insecurity. For example, CT has been found to be more effective in reducing suicide among farmers relative to the general population. However, CT did not help improving mental health of informal workers working in a hazardous work setting, while free health insurance did lower stress. In a sample of unemployed refugees, though positive, effectiveness of CT has been lower compared to workfare intervention among males relative to females. In the farmer and the refugee settings, a source of mental distress was income uncertainty, and so CT could improve their psychological well-being. In addition, males were unable to use time productively in the refugee settings, unlike the females who were involved in day-to-day household activities. So, providing employment was associated with non-pecuniary benefits by greater improvement in mental health of male participants relative to equivalent CT. However, in the context of informal workers, there was no income uncertainty as they were under wage employment. Rather, health hazard and chances of meeting with an accident was worrisome, and hence free health was effective in improving their mental health. Similarly, a study that compared CT with psychotherapy found that CT reduced depression and stress, but no significant effects were observed for psychotherapy. This contrasts with existing research which has found strong positive effects of psychotherapy in reducing depression particularly [113, 114]. The sample population for the above studies with psychotherapy as an intervention were different as for one of them it consisted of women participants with a history of sexual violence. It is likely that unlike the sample population from other psychotherapy studies, women with past experiences of sexual violence were traumatized, and hence psychotherapy was effective in improving mental health.

A second general inference that we draw form the results about the effects of an intervention is based upon the condition that the interventions place upon the beneficiaries. For example, a CCT worsened mental health of female participants relative to male, for whom the effects were positive, because the necessary compliance conditions for the CCT disproportionally burdened the females. Similarly, participants of microfinance intervention have reported stress associated with the repayment of microloans, even when the loan featured limited liability. On the contrary, a variant of microfinance, where instead of money livestock was loaned, reduced depression and PTSD among participating women. Since cash loans being liquid can be easily channeled into unproductive expenditure, whereas livestock being a productive asset demands lesser compliance conditions to repay, and hence are less worrisome.

A third inference that we draw from our review is associated with the process of delivery of the benefits from the intervention. If the benefits of an intervention are delivered using a mechanism that stigmatizes the beneficiary, this can adversely affect the mental health. Evidence of higher depression among recipients of CT social assistance program in China has been found as CT benefits are made public which invites shame and stigma for the beneficiaries.

One of the major contributions of our research is to identify gaps in the literature and opportunities for future research. While interactions with the PWLE and literature search informed us that several financial schemes exist, we did not find an evaluation of such schemes which included the mental health dimension. Examples of such schemes in India include: quarterly CT to farmers, Social Health insurance schemes, Rural informal sector employee life and disability insurance, and Pension insurance. Other than studies with experimental designs, few studies have evaluated the effectiveness of existing insurance interventions on dimensions of mental health. It was rather surprising that we did not find any study that evaluated effectiveness of crop insurance and price support insurance on mental health of agrarian workers. While Tafere et al. (2019) evaluated the livestock insurance in Ethiopia, the primary outcome was subjective well-being rather than psychological well-being. We found a sizeable number of articles that evaluated the effectiveness of pension insurance and social health insurance on mental health in China. This was possibly because data on mental health of beneficiaries was collected. Therefore, we strongly believe that one of the largest impediments to the existence of evidence based mental health research is the lack of mental health data.

Our study had several limitations too. Firstly, we only considered resources available in English for our review, so the possibility that we might have missed relevant articles in other languages remains. Secondly, though we had planned to include farmers and farm workers as PWLE from Kenya, Uganda, and Sri Lanka in our study, due to the surge in COVID-19 infections worldwide and internal economic instability in Sri Lanka, travel to these countries had to be cancelled. We believe that involving PWLE from these countries would have strengthened our research design further.

We would like to mention that not all PWLE we engaged, particularly the farmers and workers, were experiencing a mental health condition. While for many it was quite apparent during the conversation that there were symptoms of mental illnesses, often a condition had not been clinically diagnosed. Nonetheless, irrespective of the state of their mental health, contribution of each PWLE was significant for improving our research design. Therefore, we also included studies that used self-reported indicators of mental health rather than limiting our review to studies that have used clinical measures of mental health used for diagnosing mental illness.

## Recommendations for Policy and Practice

Of all the interventions reviewed, we would recommend non-contributory pensions to be one of the strongest forms of insurance intervention that has the potential to address mental illnesses among the rural working population. Among the research studies that evaluated the non-contributory pension, we found consistent positive effects of non-contributory pension on the mental health of beneficiaries or the enrollees across different countries and context, at least in the short run. This further strengthen the external validity of positive effects of non-contributory pensions.

Our next recommendation would be contributory pensions and health insurance interventions, as they have been found to have desirable positive effects on mental health of the beneficiaries in several studies. However, this recommendation is not without reservation as almost all studies except one that evaluated the effectiveness of the above two interventions, were based in China, which raises doubts about their replicability in other LMICs. This calls for evaluation of social insurance schemes for the informal workers in India which include programs like Ayushman Bharat National Health Protection Scheme, Atal Pension Scheme, future benefits under e-shram scheme and similar programs in other LMICs.

Workfare intervention has some compelling evidence towards improving mental health of beneficiaries. But the evidence comes from few specific settings, the results from which might not get translated to an another setting. Therefore, limited but encouraging evidence from workfare intervention require further research to establish the external validity of the effectiveness of workfare intervention beyond the chosen experimental settings.

CT and microcredit interventions have produced contrasting evidence in terms of their effectiveness on mental health of beneficiaries. We found that the design and the delivery mechanism of the CT largely influenced the effectiveness of the CT on the mental health of the beneficiaries. Under a CCT, if the designed compliance condition to be eligible to receive the benefits are stringent, it may adversely affect the mental health of the individual expected to adhere to the prescribed conditions. Similarly, the benefits of a CT must be delivered in a non-stigmatizing manner so that the recipients do not suffer from the perceived-stigma, which otherwise can worsen mental health. Therefore, we recommend that the CT must be used with caution with particular attention being paid to its design. Similarly, microcredit interventions must not be considered a silver bullet as they can have ambiguous effect on the beneficiaries which is highly contextual.

## Recommendations for Research

Our major recommendation for future research would be to focus on a long-term strategy to evaluate the interventions on the domain of mental health in addition to existing economic parameters. Evidence does not suggest that economic and psychological well-being following a policy intervention necessarily co-move in the same direction. This implies that not taking mental health effects into account could be underestimating or overestimating the overall benefits of the policy intervention. This calls for introducing mental health variables in the existing health databases. One of the largest impediments towards understanding the effectiveness of mental health research is the lack of mental health data in LMICs. There are already global programs like Demographic and Health Surveys (DHS) which collect health data in various countries. Programs like DHS may consider collecting mental health data too. Similar initiatives at national level can be introduced, for example, National Family and Health Survey data collection in India might consider giving serious thought to the collection of mental health data in addition to physical health variables. Moreover, mental health variables should be collected using clinically validated tools as much as possible, to ensure robust evaluation. An initiative towards evidence-based research on mental health policies would be an investment for LMICs to inform scientific and evidence-based research, practice, and policies in the domain of mental health.

## Conclusions

We reviewed the existing evidence on the effectiveness of several insurance interventions among the rural agrarian working population in LMICs. Our primary interventions included non-contributory pensions, contributory pensions, contributory and non-contributory health insurance, cash transfer, workfare, and microcredit. We recommend non-contributory pension insurance to be the most effective in improving the mental health of the beneficiaries across different countries and in various context. The recommendations regarding the rest of the interventions are not without reservation on grounds of external validity or mixed results, which indicates that generalization of the results demand further research. Moreover, we highly recommend that global health organizations and LMICs should focus on evidence based mental health research and strengthen the mental health data collection to inform evidence based mental health research, practices and policies.

## Data Availability

All data produced in the present work are contained in the manuscript.

## Acknowledgement

This research was commissioned by the Wellcome Trust. For the purpose of open access, Wellcome has applied a CC BY public copyright licence to any Author Accepted Manuscript version arising from this submission We are thankful to Binoy Majumdar, Shubham Mishra, Gauri Kulkarni, Viraj Mulik and Maharnab Naha for their excellent research assistance.

## Appendix

**Table A1:**
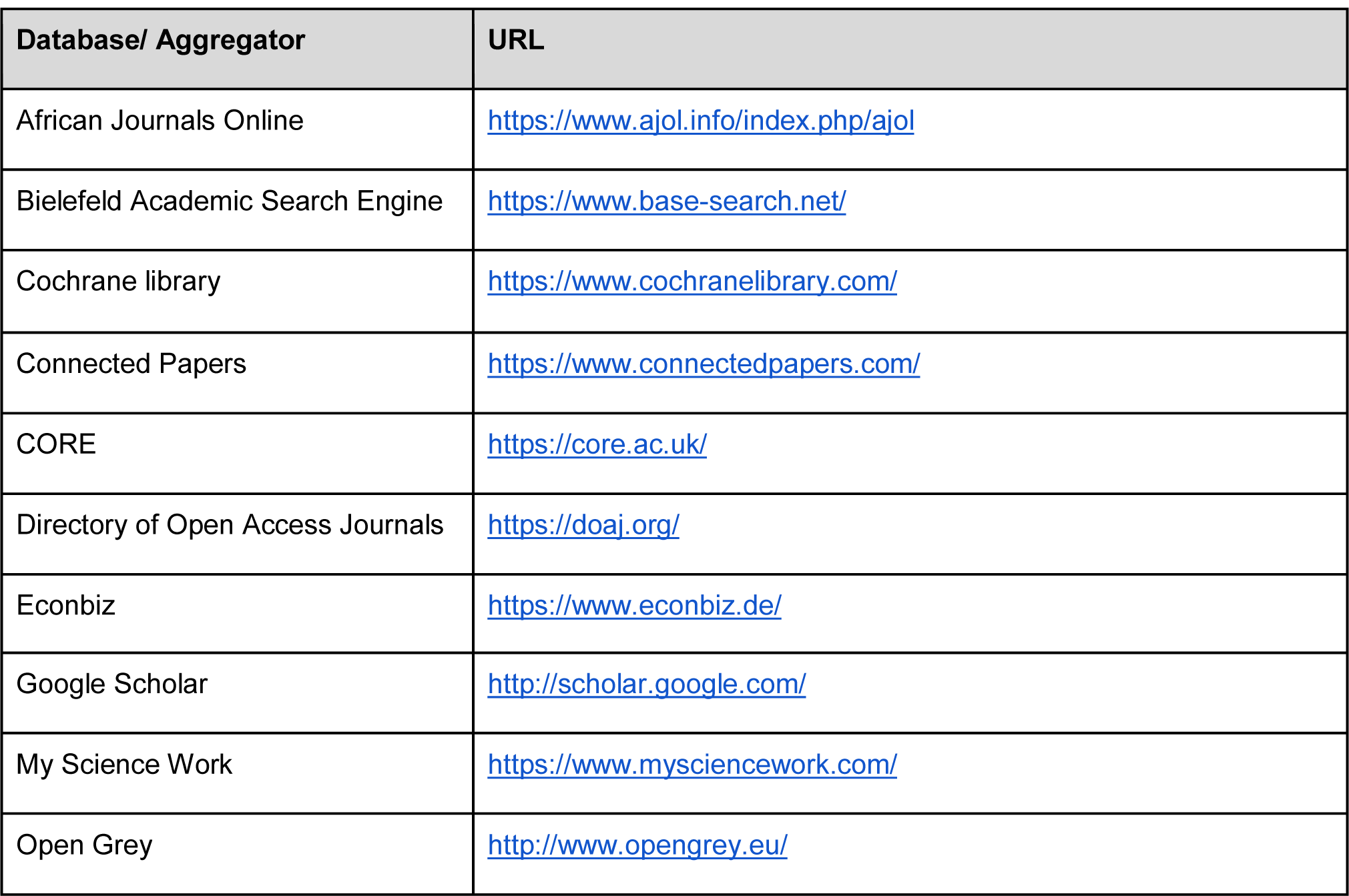
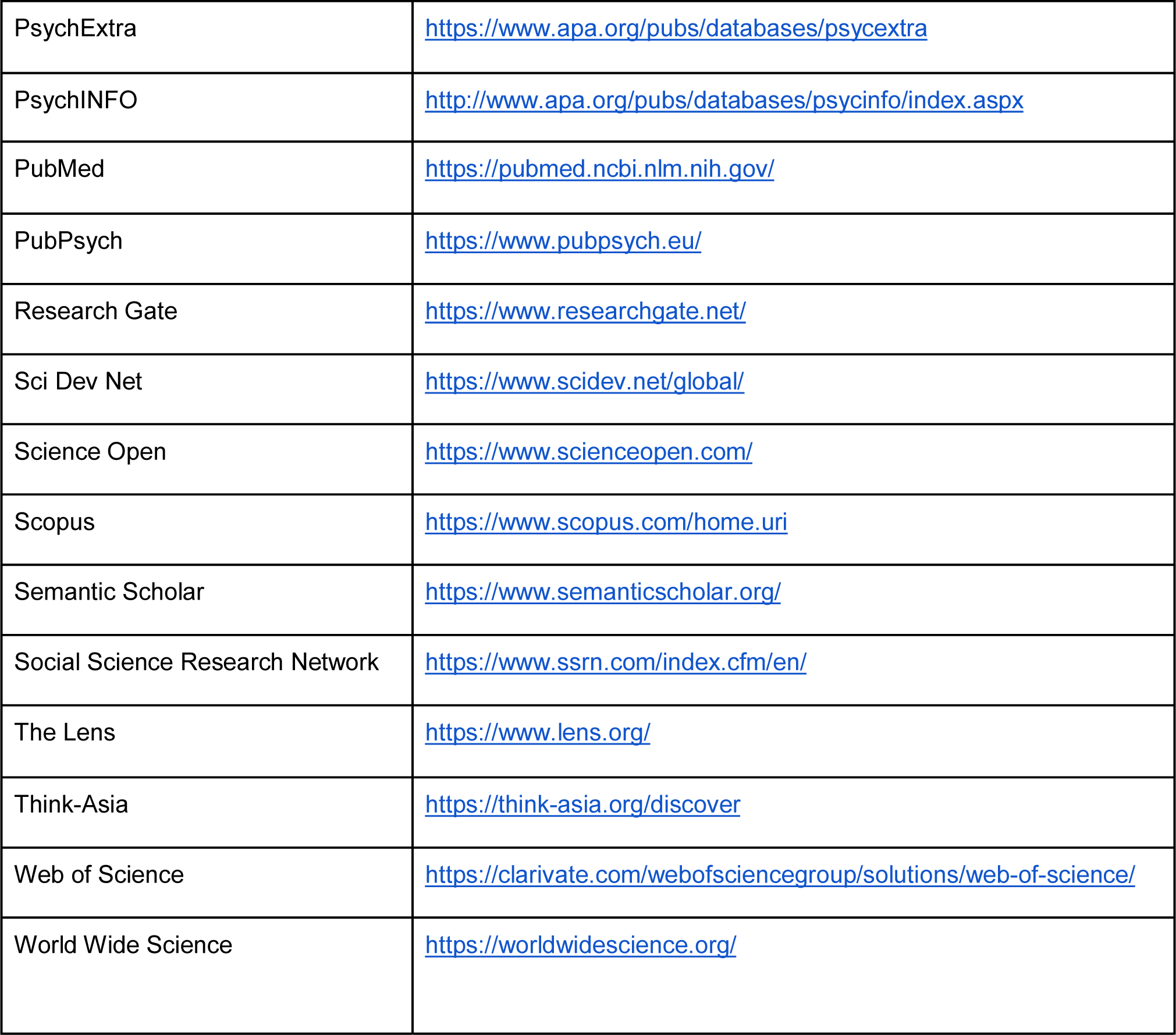
Databases and aggregators shortlisted from Wikipedia used for keyword searches (https://en.wikipedia.org/wiki/List_of_academic_databases_and_search_engines)

### Array A2: Keywords used for literature search

“Access to higher education”, “Alcoholism”, “ASHA worker”, “Awareness of mental illness”, “Banking penetration”, “Behavioural issues”, “Big hit to the prices of farm produce in the market during covid pandemic”, “Cash versus bank transfer”, “Cattle rearing”, “Climate shocks”, “Collateral for bank loan”, “Commercial crops”, “Compensation for low yield”, “Compulsory crop insurance”, “Conditions for getting free loans by Society”, “Co-operative Society”, “Corrupt system”, “Counselling”, “COVID relief transfer”, “Crop diversification”, “Crop insurance”, “Crop loss”, “Crop rotation”, “Customs”, “Cyclones and storms”, “Debt burden”, “Debt trap”, “Demographic and Family Health Survey”, “Depression”, “Drought”, “Drug abuse”, “Duplication of efforts”, “Effective social safety nets”, “Emergency loan”, “Facilities / treatment mental illness”, “Farmer Suicides”, “Farming as the only income source for the farmers who own less than two acres of land”, “Female participation”, “Fertiliser subsidy”, “Food for work”, “Fragmentation”, “Free seeds and fertilizers”, “Freedom women have at home to make important decisions related to farming”, “Gold mortgage”, “Government survey”, “Groundwater depletion”, “Hallucinations”, “Health and Wellness Centre”, “Heavy expenditures on farming till the harvest”, “Hesitancy of approaching for help”, “High labour cost”, “Hope for next year”, “Helplessness felt by farmers when crops don’t come up well and get unfair prices for it - no other ways they find to come out of this”, “Improved seed variety”, “Inability to repay loans”, “Informal credit”, “Insomnia”, “Integrated farming”, “Internal cultivation”, “Irrational expenditure”, “Kapas (Cotton) farming in Vidarbha”, “Labour cost”, “Lack of government funds”, “Land ownership”, “Lease land”, “Loan waiver”, “Low interest loans”, “Market price fluctuation”, “Mechanization of agriculture”, “Media glorification of suicides”, “Mental distress”, “Mental Health Clinic”, “Mental pressure and tense mind because of disturbed weather and less/no rainfall”, “MGNREGA”, “Microfinance”, “Middlemen”, “Migration”, “Multi-factorial etiology”, “National Income Dynamics Study”, “Natural disaster”, “Negative thoughts”, “No compensations by the government on damage”, “No expected revenue at least to recover investment in agriculture”, “No one-time payments when sold to the government at MSP”, “Nuksan Bharpai by the government (compensation)”, “Organic Farming”, “Partition of the farmland”, “Patriarchy”, “Pawning of jewellery”, “Peace of mind”, “Peer support system”, “Peer support volunteers”, “Pest infection”, “Physical disability”, “Pradhan Mantri Kisan Samman Nidhi Scheme”, “Price volatility”, “Prolonged sadness”, “Psychological autopsy”, “Relief / Rehabilitation”, “Rise in the costs of seeds, fertilizers, pesticides supplied by the government”, “Role of Gram Sevaks in MGNREGA”, “Sanstha in the village”, “Seasonal employment”, “Second job”, “Self Help Groups *(Bachat Gat) /* Microcredit”, “Self help groups”, “Selling farmland because of mental pressures and less farming income”, “Severe mental disorder”, “Shift towards other income sources because of less farming income”, “Small and medium farmers”, “Social protection”, “Society for farmers”, “Soil health card”, “Soil quality”, “Status quo bias towards farming”, “Steady income”, “Stigma/ taboo of mental illness”, “Strained family relations”, “Subsidies by the government for farming : requirements and possibilities”, “Subsidized health care”, “Sudden irritation”, “Suicidal ideation”, “Suicide attempts”, “Suicide compensation as perverse incentive”, “Unavailability of work at MGNREGA”, “Uncertain weather (rains)”, “Unproductive expenditure”, “Water management for farming”, “Water scarcity and stress”, “Ways that farmers follow to overcome the tensions”, “Wild animal attacks”, “Women farmers”, “Women reservation”, “Worries about getting good prices for the farm produce”

**Table A3:** Summary of Evidence based Studies

| Author(Year) | Country | Type of Intervention | Name of Intervention | Sample and size | Study design / Data | Psychological variables | Measurement tool | Findings |
| --- | --- | --- | --- | --- | --- | --- | --- | --- |
| Haushofer & Shapiro (2016) | Kenya | UCT | NA | 1008 rural poor households in Rarieda district | RCT | Psychological Well-being, Depression, Worry, Stress, Happiness, Life Satisfaction, Cortisol levels | CES-D, Cohen's Perceived Stress Scale, World Value Survey, Cortisol levels | CT has a positive effect on psychological well-being. |
| Haushofer & Shapiro (2018) | Kenya | UCT | NA | Long-term follow-up study of Haushofer and Shapiro (2016). | RCT | Psychological Well-being, Depression, Worry, Stress, Happiness, Life Satisfaction, Cortisol levels | CES-D, Cohen's Perceived Stress Scale, World Value Survey, Cortisol levels | Positive effect of CT on mental health persist over the long term. |
| Christian et al. (2019) | Indonesia | CCT | Program Keluarga Harapan | Farmers and rural poor from 80000 villages in Indonesia | PODES census data set (years 2000, 2003, 2005, 2011 and 2014) and | Suicides, Depression | CES-D (10-item) | CT prevents suicides from negative income shocks. |
|  |  |  |  |  | RCT |  |  |  |
| Haushofer et al. (2020b) | Kenya | UCT or Psychotherapy or both | NA | 5756 individuals from rural low-income households | RCT | Psychological well-being index, Stress, Happiness, Life satisfaction, Disability | GHQ-12, Cohen Perceived Stress Scale - 4, World Value Survey, WHODAS | CT was more effective in reducing stress and depression as compared to psychotherapy |
| Kilburn et al. (2016) | Kenya | UCT | Cash Transfer for Orphans and Vulnerable Children (CT-OVC) | 1960 youth aged between 15 and 24 years | Quasi-experiment | Depression, hope | CES-D (10 item) | CT aids in reducing depression, especially among men. The effects are not significant on female participants. |
| Hidrobo et al. (2020) | Mali | UCT with voluntary information sessions | Filets Sociaux (Jigisemejiri) | 2262 individuals from poor households | Cluster RCT | Worry, Stress/Anxiety | Cohen's Perceived Stress Scale | CT reduces anxiety. |
| Prencipe et al. (2021) | Tanzania | Social Safety Net- UCT and CCT along | Productive Social Safety Net (PSSN) | 880 individuals from the poorest 10% of | Cluster RCT | Depression, Social Support | CES-D (10 items); Multidimension | CCT reduces depressive symptoms |
|  |  | with Public Works Program |  | the population |  |  | al Scale of Perceived Social Support (MSPSS-8 item) | among male participants, but females report no reduction in depressive symptoms. |
| Hussam et al. (2021) | Bangladesh | Employment / UCT | NA | 745 adults aged between 18 and 45 years in Rohingya refugee camps | RCT | Depression, Stress, Life satisfaction, Locus of control | PHQ-9, Cohen's Perceived Stress Scale, Diener's Satisfaction With Life Scale, Rotter's thirteen-question Locus of Control Instrument | Employment / workfare has higher positive impact in terms of improving mental health among participants than cash transfers. This difference in the effects was evident among males, but not among females. |
| Haushofer et al. (2020a) | Kenya | UCT / Free Health Insurance | NA | 789 informal workers in hazardous | RCT | Subjective well-being index, | Cohen's Perceived Stress Scale | Health insurance reduces self- |
|  |  |  |  | conditions with minimal safety equipment |  | Perceived stress, Optimism, Self-esteem, Depression, Internal locus of control, Life-satisfaction, Happiness |  | reported stress, relative to receiving cash transfers. However depression is unaffected by health insurance. |
| Qi and Wu (2018) | China | Social Safety Net- UCT and CCT along with Public Works Program | Dibao | 34000 adults over the age of 16 years (from 16000 households) | China Family Panel Study Survey data investigated by the Institute of Social Science Survey at Peking University (2010, 2012, 2014 waves) | Self-satisfaction, Emotional status indicators | Self- reported indicators | Receiving income assistance through cash transfers is associated with stigma. |
| Wu et al. (2021) | China | Social Safety Net- UCT and CCT along with Public Works Program | Dibao | 4192 youth between the ages of 16 and 24 years (3345 households) | China Family Panel Study (2012 wave) | Depression | CES-D (20 items) | Receiving income assistance through cash transfers is associated with stigma. |
| Blattman et al.<br>(2014) | Uganda | Cash Grant | Youth Opportunities Program (YOP) | 11278 poor unemployed young adults between 16 and 35 years of age (535 groups) | RCT | Alienation and aggression (Participation and engagement, Social integration, Depression and distress symptoms, Aggression and hostility); Subjective-well-being (Current subjective well-being, Future subjective well-being) | Self-reported indicators | Cash grants have no significant effects on depression or stress in the short term. |
| Green et al.<br>(2016) | Uganda | Multiple interventions -<br>(i) business skills training<br>(ii) start-up grant (iii) | Women's Income Generating Support (WINGS) | 1800 young women between 14 and 30 years of age from 120 villages in | RCT | Depression | Acholi Psychosocial Assessment Instrument (APAI - 35-item) locally | Cash grants have no significant effects on mental health variables |
|  |  | business advice and encouragement to use the grant for business development. |  | post conflict settings |  |  | developed in Northern Uganda | among women. |
| Blattman et al. (2020) | Uganda | Cash Grant | Youth Opportunities Program (YOP) | 11278 poor unemployed young adults between 16 and 35 years of age (535 groups) | RCT | Alienation and aggression (Participation and engagement, Social integration, Depression and distress symptoms, Aggression and hostility); Subjective-well-being (Current subjective well-being, Future subjective well-being) | Self-reported indicators | Even in the long term, cash grants have no significant effects on depression or stress |
| Salinas-Rodríguez et al. (2014) | Mexico | Non-contributory Social Pension | Adultos Mayores | 5270 rural elderly aged 70 years and above | Quasi-experiment | Depression, Autonomy, Empowerment | Geriatric Depression Scale (GDS) | Non-contributory social pension helps reduce depression. |
| Galiani et al. (2016) | Mexico | Non-contributory Social Pension | Adultos Mayores | 3556 individuals from households with at least one adult between 66 and 74 years of age | Quasi-experiment | Depression | Geriatric Depression Scale (GDS) | Non-contributory social pension helps reduce depression. |
| Bando et al. (2017) | Peru | Non-contributory Social Pension | Pension 65 | 3342 elderly adults from 2584 households, between the ages of 65 and 80, marginally below and above the poverty index | Primary survey conducted by National Institute of Statistics (Instituto Nacional de Estadística e Informática or INEI) | Depression | Geriatric Depression Scale (GDS) | Non-contributory social pension helps reduce depression. |
| Alzua et al.<br>(2020) | Nigeria | Non-contributory Social Pension | NA | 6326 rural elders above 65 years of age with monthly income less than NGN 3000 | Cluster RCT | Depression and Self-esteem | Geriatric Depression Scale (GDS), Self-reported indicators | Non-contributory social pension helps reduce depression. |
| Bando et al.<br>(2022) | Paraguay | Non-contributory Social Pension | Pensión Alimentaria | 2473 poor adults above 65 years of age | RCT | Depression, Satisfaction, Empowerment, Contribution, Self-worth, Well-being | Geriatric Depression Scale (GDS), Self-reported indicators | Non-contributory social pension helps reduce depression. |
| Chen et al.<br>(2019) | China | Social Pension Insurance - Contributory but Subsidized | New Rural Social Pension Scheme | 8636 rural individuals above 45 years of age | China Family Panel Study survey (2010, 2012 waves) | Depression | CES-D (20 items) | Contributory social pensions reduce depression among elderly in rural areas. |
| Ma & Oshio<br>(2020) | China | Social Health insurance and Pension Insurance | New Rural Social Pension Insurance (NRSPI), New Rural | 32808 rural adults from Hukou population above 45 | China Health and Retirement Longitudinal Study (2011, | Present and Prospective Mental Health | Self-reported indicators | Contributory social pensions reduce depression |
|  |  |  | Cooperative Medical Scheme (NRCMS) | years of age | 2013, 2015 waves) |  |  | among elderly in rural areas. However social health insurance has no significant effects on psychological well-being. |
| Pan et al. (2021) | China | Social Pension Insurance - Contributory but Subsidized | New Rural Social Pension Scheme | 21122 rural individuals above 45 years of age | China Health and Retirement Longitudinal Study (2015, 2018 waves) | Depression | CES-D (10 items) | Contributory social pensions reduce depression among elderly in rural areas. |
| Wang & Zheng (2021) | China | Social Pension Insurance - Contributory but Subsidized | New Rural Society Pension Insurance | 1362 rural households with elders above 60 years of age | China Health and Retirement Longitudinal Study (2011, 2013 waves) | Depression | CES-D (10 items) | Contributory social pensions reduce depression among elderly in rural areas. Effects observed among elderly females, but not among males. |
| He et al. (2021) | China | Social Pension Insurance - Contributory but Subsidized | Basic Pension system for urban workers (UW), Pension system of Public institutions (PI), Basic Pension system for urban and rural residents (URR), New Rural Pension | 6687 elderly individuals 60 years old and above | Chinese Longitudinal Aging Social Survey (2014) | Depression | CES-D | Social pensions are associated with lower depression scores, but causality needs to be validated. |
| Lloyd-Sherlock & Agarwal (2014) | South Africa | Pension (may include contributory and non-contributory) | Old Age Grant | Approximately 2000 black South African elderly women aged 60 years and above, men aged 63 years and above | WHO Study of global AGEing and adult health | Depression, Anxiety | Self-reported indicators | Effects of pension on depression and anxiety are not significant. |
| Wang et al.<br>(2009) | China | Social Health Insurance - Community based Contributory but Subsidized | Rural Mutual Health Care (RMHC) - Experiment | 7025 individuals aged 15 years and above from 1925 households | Quasi-experiment | Depression, Anxiety | Categorical Rating Scale (CRS), EQ-5D | Social health insurance reduces depression. Effects were most significant among people older than 55 years, and ill during the time of intervention. |
| Tian et al.<br>(2012) | China | Health Insurance | NA | 4079 individuals from households in Lanzhou City and Baiyin City in Gansu province | Clustered sampling primary survey | Depression | CES-D | Social health insurance reduces depression. |
| Sun & Lyu<br>(2020) | China | Social Health Insurance - Contributory but Subsidized | New Rural Cooperative Medical Scheme (NRCMS) | 2093 rural adults aged 45 years and above | China Family Panel Study (2016 wave) | Depression | whether or not they had felt uncomfortable in the previous 2 weeks, Frequency at which they had felt sad over the previous week | Social health insurance reduces depression. Low-income individuals benefit the most in terms of mental health improvements. |
| Sun et al. (2021) | China | Social Health Insurance - Contributory but Subsidized | Urban and Rural Resident Basic Medical Insurance (URRBMI) | 2198 adults aged 45 years and above | China Health and Retirement Longitudinal Study (2018 wave) | Depression | CES-D (10-item) | Social health insurance reduces depression. Higher effects observed among males less than 65 years of age. |
| Shieh et al. (2021) | China | Social Health Insurance | Catastrophic Medical Insurance (CMI) | 4727 old adults aged 60 years and above | Chinese Longitudinal Healthy Longevity Survey (conducted by Peking University and China Scientific Research Centre on | Index of Positive well-being, Index of negative well-being | Self-reported indicators | Catastrophic medical insurance reduces anxiety and depression among elderly. |
|  |  |  |  |  | Ageing) (2011-12 and 2014 waves) |  |  |  |
| Yang & Hu (2022) | China | Social Health Insurance | Urban Employee Insurance (UEI), Urban Resident Insurance (URI), New Rural Cooperative Medical Scheme (NCMS) | 13166 old adults aged 60 years and above | China Health and Retirement Longitudinal Study (2011, 2013 and 2015 waves) | Depression | CES-D (10 items) | Social health insurance is associated with lower depression scores, even with catastrophic medical expenditure. |
| Ravi & Engler (2015) | India | Public Workfare Programme | National Rural Employment Guarantee Scheme | 1064 ultra poor households in Medak district of Andhra Pradesh | Primary survey | Felt Worried/Tense d/Anxious for one month in previous year | Self-reported indicators | Workfare helps reduce anxiety. |
| Tsaneva & Balakrishnan (2019) | India | Public Workfare Programme | National Rural Employment Guarantee Scheme | Approx. 3905 (136 districts) recovered from supplementary appendix | WHO Study of global AGEing and adult health | Depression, Stress, Psychological Well-being | ICD-10, Self-reported indicators | Workfare reduces depression among rural women. |
| Bhanot et al.<br>(2018) | Kenya | Employment /<br>UCT | NA | 432 individuals<br>from low<br>income<br>households in<br>Kawangware<br>settlement in<br>Nairobi | Randomized<br>experiment | Mental Health,<br>Well-being<br>index | PANAS (6<br>item), Self-reported<br>indicators | Working for<br>benefits has<br>much more<br>positive impact<br>on<br>psychological<br>well-being,<br>than receiving<br>benefits<br>without<br>working. |
| Pailler &<br>Tsaneva<br>(2018) | India | Public<br>Workfare<br>Programme | National Rural<br>Employment<br>Guarantee<br>Scheme | 16227<br>working-age<br>adults between<br>18 years and<br>60 years of<br>age | World Health<br>Survey (WHS)<br>and the WHO<br>Study of global<br>AGEing and<br>adult health<br>(SAGE) | Depression,<br>Stress,<br>Psychological<br>Well-being | Self-reported<br>indicators | Workfare helps<br>mitigate the<br>negative<br>effects of<br>climate change<br>on mental<br>health of<br>participants. |
| Mohindra et al.<br>(2008) | India | Microfinance | Self-help<br>Groups -<br>Kudumbrasree | 928 women<br>between 18<br>years and 59<br>years<br>(excluding the<br>Paniya group)<br>in Kottathara, | Primary<br>household<br>survey | Frequency of<br>disturbance in<br>mental peace;<br>Life<br>satisfaction | Self-reported<br>indicators | Microfinance<br>reduces<br>emotional<br>stress among<br>female<br>farmers. |
|  |  |  |  | Wayanad district in Kerala |  |  |  |  |
| Hamad & Fernald (2015) | Peru | Microfinance | Prisma (NGO) microcredit intervention | 1593 women in Pucallpa who are PRISMA clients | Primary cross-sectional survey | Depression, Perceived Control on life circumstances, Perceived social support | CES-D (20 items, range 0-60); Duke-UNC Functional Social Support Questionnaire (11 items, range 0–55) | Microcredit reduces depression among women borrowers. |
| Bass et al. (2016) | Congo | Microfinance | Village Savings and Loans Associations (VSLA) | 301 female sexual violence survivors | RCT | Depression, Anxiety, Post-traumatic stress | Hopkins Symptom Checklist-25 (HSCL-25), Harvard Trauma Questionnaire (HTQ), Self-reported indicator | No reduction in symptoms of depression, anxiety, or PTSD. |
| Glass et al. (2014) | Congo | Microfinance - Livestock intervention | Pigs for Peace (PFP) | 649 rural women in settings of | Randomized Community Trial | Post-traumatic stress syndrome | Harvard Trauma Questionnaire | Microcredit in the form of animal assets |
|  |  |  |  | conflict and violence |  | (PTSD), Depression | (HTQ); Hopkins Symptom Checklist (HSCL) | reduces depression and PTSD among women. |
| Ahmed et al. (2001) | Bangladesh | Microfinance | Building Resources Across Communities (BRAC NGO) microcredit intervention | 2075 married women between 15 years and 55 years from poor households | Primary cross-sectional survey | Emotional stress, Depression | Self-reported indicators | Microcredit is associated with higher emotional stress among women. |
| Bhuiyan & Ivlevs (2019) | Bangladesh | Microfinance | NA | 1430 households in three villages in Tangail district, Bangladesh | Village census survey | Subjective well-being, Negative affect (worry and depression), Positive affect (happiness) | Self-reported indicators | Microcredit is associated with higher emotional stress among women. |
| Fernald et al. (2008) | South Africa | Microfinance | NA | 787 marginally rejected loan applicants of one of the largest urban micro-lender | RCT | Stress, Depression | CES-D (20 items, range 0-60); Cohen's Perceived Stress Scale (10 items, range 0-40), | Access to microcredit decreases depression but is associated with elevated stress among borrowers. |
| Hatcher et al. (2020) | Kenya | Multiple interventions -<br>(i) a low-cost micro-irrigation water pump (ii) training on sustainable farming practices and financial management (iii) loan program of vouchers (worth □\$150 USD) | Shamba Maisha | 54 HIV positive farmers | In-depth interviews | Stress and Anxiety | Qualitative Description | Reduction in symptoms of anxiety and depression. |
| Liang et al. (2014) | China | Social Security Policies - Land Acquisition Compensation Policy, Medical Insurance Policy, Pension | New Rural Cooperative Medical Care Insurance | 1236 landless farmers | Household survey | Health-related quality of life (HRQOL) on 5 dimensions - Mobility, Self-care ability, Daily activities, Pain/Discomfor | EQ-5D | Social security policies are associated with lower levels of depression and anxiety among |
|  |  | Insurance Policy, Employment Security Policy, Basic Livelihood Guarantee Policy, Housing Compensation Policy |  |  |  | t, and Anxiety/Depression |  | landless farmers. |
| Banerjee et al. (2015) | Ethiopia, Ghana, Honduras, India, Pakistan, Peru | Multiple interventions | Graduation Programme | 10495 ultra poor households | RCT and Cluster RCT | Mental Health Index, Stress Index | Self-reported indicators | No significant improvement in psychological well-being of participants. |
| Burmester et al. (2015) | Dominica | Living Wage Intervention and Key work place improvements | NA | 204 apparel factory workers | Quasi-experiment | Depression | CES-D | Living wage intervention is associated with lower depression scores. |
| Richardson et al. (2018) | India | Subsidized Day Care | NA | 3041 rural women in low income<br>settings | Cluster RCT | Mental distress | GHQ-12 | Access to low-cost daycare resulted in<br>modest<br>reduction of<br>mental distress<br>among rural<br>women. |

### A4 Suggestion based studies

In this section we present a review of literature that provides recommendations or suggestions that would reduce agrarian distress rather than any evidence-base as was derived from original research papers. Except for Kandlur et al. (2022) that measured mental distress among farmers through anxiety, depression, distress and psychological morbidity as the mental health variables, all studies that reviewed in these studies equated agrarian distress with agrarian suicides. Moreover, all our reviewed studies were from India except from Knipe et al. (2014), which was about suicides in rural Sri Lanka and Murray and Mahat (2017), which was conducted among the bonded labor in Nepal.

Price insurance against low market prices in the form of Minimum Support Prices had been strongly advocated as a potential insurance intervention to reduce agrarian distress and farmers suicides [81, 83-97]. Crop insurance had been suggested as one of the most important insurance intervention that can potentially mitigate suicides by extending financial security in the event of crop loss [83-86,88,90-92,94,95,98-100,101-103]. While Parida et al. (2018) advocated compulsory crop insurance for all farmers, Bhojanna and Raturi (2012) suggested that corporates could bear the cost of crop insurance premium under Corporate Social Responsibility (CSR), which would save farmer’s lives. Sulaiman and Murigi (2018) in a hypothetical cost benefit analysis of crop insurance estimated the number of farmers’ lives that can be saved in a year by crop insurance. However, Van de Meenderdonk (2020) found in an ethnographic study in India that farmers reported crop insurance to be ineffective in reducing their distress. Rao et al. (2017) suggested designing comprehensive crop insurance such that it covered loss at every stage of the crop cycle rather than only at harvest.

Alternative livelihoods or off-farm employment similar to NREGS could potentially reduce farmers’ suicides by reducing dependence upon highly volatile farm income [85,90,95,96,98,102,105]. Knipe et al. (2014) suggested that implementation of the minimum daily wages for laborers could reduce rural suicides in Sri Lanka. Parida et al. (2018) suggested on similar lines that higher agricultural wage income could potentially reduce agrarian distress. Few other suggestions to reduce suicides included provision of health insurance and pension insurance for farmers [95, 98].

Suggestions had been also made that access to formal credit at zero or low interest rate could also reduce farmer’s suicides [84,85,91,92,97,98,100,106]. Credit access from microfinance could also contribute in reducing agrarian distress [82,88,94]. Murray and Mahat (2017) in a study among bonded manual labor, which is a form of modern slavery in Nepal, suggested that the incidence of anxiety, depression and PTSD among the bonded labor could be potentially reduced by their participation in microfinance. However, Taylor (2011) argued that microfinance was not a solution to agrarian distress citing the example of microfinance crisis in Andhra Pradesh, India.

Heavy indebtedness among the farmers had been associated with suicides. As a consequence, there had been suggestions of loan moratorium and loan waivers, which could relieve indebtedness, and hence can reduce suicides among farmers [84, 88, 89, 108, 109]. However, most authors also acknowledged that the debt relief measures offered temporary relief and generally lacked long-term effectiveness [108–110]. Kandlur et al. (2022) suggested that loan waivers are unlikely to reduce depression, anxiety or distress among farmers. Further, Ravi (2015) critically puts forward that instead of combating, loan waivers and cash supports rather aggravate the issue of farmer suicides.

**Table A5:** Summary of Suggestion based Studies

| Author (Year) | Country | Type of Intervention | Reference Population | Hypothesis |
| --- | --- | --- | --- | --- |
| Rangarajan & Dev (2019) | India | MSP | Farmers | MSP brings higher income for farmers and dispel agrarian distress |
| Guha (2012) | India | MSP |  | MSP acts as a cushion and could help mitigate farmer suicides. |
| Deshpande (2002) | India | MSP, CI | Suicide victim families | Administering MSP and CI could act as safety nets to deal with farmers' distress |
| Kennedy & Kling (2014) | India | MSP, Loan Moratorium | Indebted farmers | Loan moratoriums could relieve distressed farmers and reduce suicide among them. |
| Knipe et al. (2014) | Sri Lanka | MSP, Minimum Daily Wage for Labourers | farmers, daily wage labourers, and foreign migrant households, and significantly elevated in female headed households | MSP could help reduce suicides among farmers |
| Iyer (xxxx) | India | MSP, Alternative Livelihoods | Farmers | MSP or alternative livelihoods provide relief against indebtedness and reduce farmer suicides |
| Behere et al. (2021) | India | MSP, Low-interest Formal Credit | Farmers | Recurring debt is the primary reason for farmer suicide; farm loans at affordable interest loans could eliminate financial stress and thereby reduce farmer suicide. |
| Grover et al. (2016) | India | MSP, CI, Alternative Livelihoods | Agrarian community | Increase in MSP improves income; this and crop insurance might help lessen suicides among farmers. |
| Das (2016) | India | MSP, Agricultural Insurance, Low-interest Formal Credit | Agrarian community | Agriculture insurance reduces risk and thereby agrarian distress |
| Rao et al. (2017) | India | MSP, CI, Soft Interest Farm Loans | Farmers | MSP, CI and soft interest loans help reduce agrarian distress |
| Manjunatha & Ramappam (2017) | India | MSP, CI, low-interest formal credit | Agrarian community | MSP and bank loans at low interest rates help reduce farmer suicides |
| Mukherjee (2009) | India | MSP, CI, alternate livelihood, low-interest formal credit | Agrarian community | Financial interventions may reduce suicides among farmers |
| Jeromi (2007) | India | MSP, CI, low-interest formal credit, loan waivers | Plantation farmers | Formal credit at low interest rates could help prevent farmer suicides |
| Raghavalu (2013) | India | MSP, CI, microfinance, moratorium on credit repayment | Agrarian community | Financial interventions may reduce suicides among farmers |
| Sravanth & Sundaram (2019) | India | MSP, CI, Microcredit through SHGs, price stabilization fund, loan waiver | Agrarian community | Loan moratorium cannot reduce farmer suicides |
| Chouhan et al. (2019) | India | MSP, CI, alternative livelihood, health insurance, pension insurance | Families of suicide victims | Wider insurance coverage can decrease farmer suicides in Madhya Pradesh |
| Meerendonk (2020) | India | CI | Indebted farmers | Crop insurance might help address agrarian distress |
| Bhojanna & Raturi (2012) | India | CI costs borne by corporates (Corporate Social Responsibility), soft rate interest loans | small & marginal farmers | Crop insurance protects farmers against financial loss and reduces agrarian distress |
| Sulaiman & Murigi (2018) | India | Graduation Program including CI | Ultra-Poor | Crop insurance protects from risk of crop loss and income loss; however cost of intervention is very high |
| Jabeen (2019) | India | CI | Agrarian community | Crop insurance helps deal with indebtedness and reduce agrarian distress |
| Parida et al. (2017) | India | Compulsory CI, alternative livelihood, higher agricultural wages, loan waiver | Agrarian community in drought and flood affected regions | Compulsory crop insurance and loan waivers could help reduce agrarian distress |
| Dongre et al. (2012) | India | CI | Rural community | Crop insurance can alleviate debt issues and thereby reduce farmer suicides |
| Kaur (2010) | India | CI, low-interest formal credit, alternative livelihood, health insurance, pension insurance | Civil societies in Punjab, villagers, suicide-affected families | Financial interventions create safety net and could reduce suicides among farmers |
| Mishra (2007) | India | Alternative livelihood | Agrarian community | Alternative livelihood like NREGS can help reduce farmer suicides |
| Taylor (2011) | India | Microfinance | Agrarian community | Unless regulated, MFI cannot help deal with farmer suicides |
| Murray & Mahat (2017) | Nepal | Microfinance through SHGs | Haruwa and Charuwa: Bonded and forced labourers | SHG participation could reduce depressive symptoms and PTSD among bonded manual labourers. |
| Sadanandan (2014) | India | Low-interest formal credit | Farmers | Low interest formal credit could help reduce farmer suicides, particularly in suicide prone states of India |
| Ravi (2015) | India | Loan waivers | Farmers, housewives | Loan waivers are not effective in countering farmer suicides |
| Nair & Menon (2007) | India | Loan moratorium | Agrarian community | Moratorium on loan payment can reduce financial burden and help reduce farmer suicides |
| Parvathamma (2016) | India | Loan moratorium | Agrarian community | Moratorium on loan payment can reduce agrarian distress |
| Sangalad (2012) | India | Loan moratorium | Suicide victims' and control families | Temporary relief in the form of loan moratorium won't help reduce farmer suicides |
| Kandlur et al. (2022) | India | Loan waivers | Farmers who are members of cooperatives | Loan waivers need not reduce the distress of all farmers |

MSP: Minimum Support Price; CI: Crop insurance

